# Dysregulation of circulating protease activity in Covid-19-associated superinfection

**DOI:** 10.1101/2021.10.20.21265115

**Authors:** Fernando Dos Santos, Joyce B. Li, Nathalia Juocys, Rafi Mazor, Laura Beretta, Nicole G. Coufal, Michael T.Y. Lam, Mazen F. Odish, Maria C. Irigoyen, Anthony J. O’Donoghue, Federico Aletti, Erik B. Kistler

## Abstract

Infection by SARS-CoV-2 and subsequent COVID-19 can cause viral sepsis and septic shock. Several complications have been observed in patients admitted to the intensive care unit (ICU) with COVID-19, one of those being bacterial superinfection. Based on prior evidence that dysregulated systemwide proteolysis is associated with death in bacterial septic shock, we investigated whether protease activity and proteolysis could be elevated in COVID-19-induced sepsis with bacterial superinfection. In particular, we sought to assess the possible implications on the regulation of protein systems, such as for instance the proteins and enzymes involved in the clotting cascade.

Blood samples collected at multiple time points during the ICU stay of four COVID-19 patients were analyzed to quantify: a) the circulating proteome and peptidome by mass spectrometry; b) plasma enzymatic activity of trypsin-like substrates and five clotting factors (plasmin, thrombin, factor VII, factor IX, factor X) by a fluorogenic assay.

Of the four patients, one was diagnosed with bacterial superinfection on day 7 after beginning of the study and later died. The other three patients all survived (ICU length-of-stay 11.25±6.55 days, hospital stay of 15.25±7.18 days). Spikes in protease activity (factor VII, trypsin-like activity) were detected on day 7 for the patient who died. Corresponding increases in the total intensity of peptides derived by hydrolysis of plasma proteins, especially of fibrinogen degradation products, and a general reduction of coagulation proteins, were measured as well. A downregulation of endogenous enzymatic inhibitors, in particular trypsin inhibitors, characterized the non-surviving patient throughout her ICU stay. Enzymatic activity was stable in the patients who survived.

Our study highlights the potential of multiomics approaches, combined with quantitative analysis of enzymatic activity, to i) shed light on proteolysis as a possible pathological mechanism in sepsis and septic shock, including COVID-19-induced sepsis; ii) provide additional insight into malfunctioning protease-mediated systems, such as the coagulation cascade; and iii) describe the progression of COVID-19 with bacterial superinfection.

## 1. Introduction

The Coronavirus disease (COVID-19) pandemic is caused by viral infection with the coronavirus SARS-CoV-2. Affecting every organ of the body following initial infection of the respiratory system, this highly contagious virus has contributed to the deaths of almost 5 million people to date. Despite the emphasis on COVID-19 research, our understanding of the disease remains incomplete and therapeutic options are limited (1-7). Frequent and fatal complications arising from SARS-CoV-2-induced sepsis include cytokine storm and coagulopathies (8,9). Less frequent (10), but not less dangerous, bacterial superinfections make the clinical treatment of COVID-19 patients in the intensive care unit (ICU) even more challenging, especially because of the lack of sound hypotheses as to the underlying pathology and interplay between viral and bacterial infection and their clinical implications (11).

We have previously shown, in both animal and human studies, that a possible pathological mechanism triggered by acute illness, including bacterial sepsis and septic shock, is dysregulated proteolysis (12-15). Major protease-mediated systems are known to be affected in critical illness, including activation of the coagulation and complement cascades. To better understand the possible relationship between systemic proteolysis occurring in shock, including COVID-19-induced septic shock and the malfunction of these protease-mediated systems, we have developed a complex, multimodal approach to assess protease activity and proteolysis in plasma, consisting of quantitative enzymatic activity assays and mass spectrometry-based multi-omics analyses of the circulating proteome and peptidome. The utilization of these techniques has shown promising results in the ability to i) estimate in vivo proteolysis in plasma as a marker of ongoing systemic proteolysis during shock (13,14) and ii) detect circulating peptidase activity (16).

The hypothesis of this study was that a complex, dynamic description of plasma proteins and the activity of some of their most important circulating enzymatic substrates can be leveraged as a tool to shed light on the progression of COVID-19 and the risk for fatal complications, especially in the presence of bacterial superinfection and diagnosis of sepsis and septic shock. The specific goals that we aimed to investigate were to assess: 1) the patterns of protease activity in the plasma of COVID-19 patients following admission to the ICU by means of quantitative analysis of enzymatic activity; 2) the relationship between alterations in protease activity over time during ICU stay and the development of bacterial superinfection; 3) the possible implications of dysregulated proteolysis on major protein systems (e.g., clotting proteins and enzymes) by analysis of plasma proteomics and peptidomics.

## 2. Materials and Methods

### 2.1 Clinical measurements

This was a prospective observational study. The University of California, San Diego Institutional Review Board (IRB) provided approval of the study conduct (IRB# 190699, Protocol #20-0006); informed consent was obtained for the collection of blood samples and de-identified use of the data. A convenience sample of four adult critically ill patients with confirmed positive test for COVID-19 by real-time reverse transcription polymerase chain reaction (RT-PCR) admitted to the Intensive Care Units (ICUs) at the University of California, San Diego Medical Center between April 01, 2020, and May 30, 2020, was enrolled in the study. De-identified demographic and prior health history data were obtained. Three healthy subjects were enrolled as a control group for enzymatic activity assays.

Standard laboratory measurements were obtained as necessary for optimal patient care by the UC San Diego Medical Center for Advanced Laboratory Medicine (CALM) laboratories as part of regular and usual care. No additional laboratory data were collected.

All non-standard laboratory assays, including protease activity assays and mass spectrometry measurements were conducted off-site in academic research laboratories at the University of California, San Diego. Patient blood samples (3-10mL) were collected in sodium heparin (BD Vacutainer, REF: 366480) tubes. To prevent coagulation, the tubes were inverted 10 times prior to transport at room temperature. Blood was processed within 4 hours of collection and kept at room temperature throughout the protocol. Polymorphprep™ was added to collected samples and centrifuged at 500g x 30 mins to separate plasma from white blood cells per manufacturer’s instructions (Progen) and transferred to new Eppendorf tubes and centrifuged at 3731 x g for 5 minutes to remove any cellular debris. The supernatant was transferred to new tubes and flash frozen in dry ice and 95% ethanol. Plasma was aliquoted and stored at -80° C for further analysis.

The clinical data acquisition and blood sample collection started on the day following protocol study consent (usually day of ICU admission) and was repeated on the following day and every other day thereafter until patient discharge or death.

### 2.2 Enzymatic activity

To test the enzymatic activity in COVID-19 plasma samples, we selected a variety of fluorogenic substrates, with a specific focus on enzymes with trypsin-like activity and clotting factors. All plasma samples were processed and analyzed simultaneously for each substrate, including healthy control plasma. The peptide sequences used in this assay were chosen as substrates for coagulation factor enzymes, tryptic enzymes in general, as well as endopeptidase activity (**Supplementary File 1**). Briefly, a 384-well plate was used to combine 15μL of plasma (1/50 final dilution in PBS, pH=7.4) with 15μL of each substrate (5μM in PBS) in triplicate wells and immediately measured for activity with a spectrophotometer (FilterMax F5 Multi-Mode, Molecular Devices) with excitation of 360 nm and emission of 460 nm. Wells were read every 2 minutes and 45 seconds for 4 hours and 50 minutes thereby generating fluorescent activity progress curves that each contain 100 data points. Fluorogenic activity was reported as relative fluorescence units per second using the slope from the progress curve where the r^2^ value was higher than 0.98. The activity was corrected by the dilution factor. Due to the large variance measured between patients, all results shown are plotted as the percentage of enzymatic activity relative to average of healthy control plasma samples on a log_2_ scale.

### 2.3 Proteomics and Peptidomics Analysis of COVID-19 Plasma

All plasma samples from the COVID-19 patients were subjected to a series of protein extraction and pretreatment steps prior to mass spectrometry. The complete protocols for plasma sample preparation, processing and data analysis steps can be found in **Supplementary File 1**. Briefly, for proteomics measurements, proteins were precipitated from the plasma samples, treated with denaturing, reducing, and alkylating reagents, and enzymatically digested with trypsin. To quantify peptides in plasma, high molecular weight proteins were first removed following addition of methanol and the remaining peptide quantified by LC-MS/MS. PEAKS Studio 8.5 (Bioinformatics Solutions, Inc.) proteomics software was used for protein identification and quantification.

For the purposes of our study, the analysis of the circulating proteome and peptidome focused on three main groups of proteins, according to their function: 1) coagulation; 2) complement system, immune response and inflammation; and 3) endogenous protease inhibitors. Each of these systems is known to be affected by acute illness, and their clinical management poses significant challenges in critically ill patients. The proteins most relevant to our analyses included: a) coagulation factors and fibrinogen for coagulation analysis; b) complement factors, vascular endothelial growth factor receptor 2 (VEGF2) and C-reactive protein for immune response and inflammation; c) serpins such as trypsin and chymotrypsin inhibitors, thrombin and plasmin inhibitors, C1 inhibitors and heparin cofactor II for endogenous enzyme inhibitors.

For proteome analysis, we used the log_2_ ratio of each individual intensity measurement to the average of all sample intensities for each protein. Heat maps for the three main protein function groups considered for analysis were generated and analyzed using the web-based visualization software Morpheus (https://software.broadinstitute.org/morpheus). Identified proteins were hierarchically clustered with the one minus Pearson’s correlation metric and average linkage method for each protein group. Columns were sorted by patient and ordered by day of sample collection.

For peptidomic analysis, the degree of overall proteolysis was estimated by the sum of all fragment intensities in a sample. Similarly, the proteolysis of a single protein was estimated by the sum of the intensities of the peptides generated by that protein. An online web tool, Peptigram (@Bioware - © University College Dublin) (17) was used to build cleavage maps representative of the number of peptides generated by a parent protein, abundance and length of each amino acid sequence, and cleavage site on the parent protein of origin.

## 3. Results

Patient characteristics are described in **Table 1**. The four patients randomly selected for analysis were comprised of three males and one female and ranged in age from 29-88 years. Reflecting the overall demographics of the COVID-19 epidemic in San Diego, three of the subjects were Hispanic. One patient, who will be referred to as “Patient 3” in the following, was diagnosed with bacterial superinfection and subsequent sepsis on day 7 and died on day 16 of her ICU stay. The remainder (Patients 1, 2 and 4) were discharged from the ICU with a mean ICU length-of-stay for all subjects of 11.25±6.55 days and mean hospital stay of 15.25±7.18 days. Body mass index (BMI) was 28.84±3.65 kg/m^2^ (range: 29.4-32.14 kg/m^2^) and self-described baseline health was either “fit” or “well”.

**Table 1.** Characteristics of COVID+ patient cohort

| Covariate | Patient 1 | Patient 2 | Patient 3 | Patient 4 |
| --- | --- | --- | --- | --- |
| BMI | 29.40 | 23.66 | 32.14 | 30.19 |
| Sex (M: male; F: female) | M | M | F | M |
| Survived (Yes / No) | Y | Y | N | Y |
| Admit to ICU from ED or Ward | Ward | ED | Ward | ED |
| ICU Length of Stay | 17 | 3 | 16 | 9 |
| Hospital Length of Stay | 24 | 8 | 18 | 11 |
| Self-described fitness level upon admission | Fit | Fit | Well | Fit |
| Modified Rankin Score on admission | No symptoms | No symptoms | No symptoms | No significant disability |
| *Treatment in ICU* |  |  |  |  |
| Required pressors | Y |  | Y | Y |
| (ARDS) diagnosis | Y |  | Y | Y |
| Presence of another infection |  |  | Y* |  |
| Antibiotics received: Azithromycin | Y | Y | Y | Y |
| Ceftriaxone | Y | Y |  | Y |
| Meropenem |  |  | Y |  |
| Vancomycin |  |  | Y | Y |
| Piperacillin/Tazobactam |  |  | Y | Y |

All patients were initially treated with azithromycin. The three surviving subjects, who did not present with a bacterial superinfection, were also treated with ceftriaxone. Patient 3 was treated after positive culture identification (*Pseudomonas aeruginosa* in urine and *Enterobacter aerogenes* pneumonia) with piperacillin/tazobactam and later, with meropenem and vancomycin, after clinical failure to respond to infection. The three more critically ill patients’ pulmonary function reflected a diagnosis of Acute Respiratory Distress Syndrome (ARDS) and these patients were intubated and placed on mechanical ventilation for at least some period of their ICU stay. These three patients also required the application of vasopressor agents to maintain hemodynamic stability.

Pertinent laboratory data are reported in **Table 2** and reflect a relatively homogeneous distribution of their hemogram as well as metabolic laboratory results (not shown) and coagulation panel. It should be noted that the outlying results in Patient 3’s white blood count (WBC) of 19.8×109/L, hemoglobin of 6.4 g/dL, and platelet count of 461 x103 μL/ml are from the final day of the patient’s hospital stay and represent her moribund condition.

**Table 2.** Main Laboratory Data

| Covariate | Patient 1 | Patient 2 | Patient 3 | Patient 4 |
| --- | --- | --- | --- | --- |
| White Blood Count (10<sup>9</sup>/l) |  |  |  |  |
| Average [SD] | 10.2 [1.5] | 8.4 [0.8] | 11.6 [4] | 7.7 [1.7] |
| (Range) | (7.6-12.3) | (7.4-9.6) | (8.1-19.8) | (5.7-10.3) |
| Hemoglobin (g/dl) |  |  |  |  |
| Average [SD] | 13.0 [1.3] | 14.1 [1.2] | 9.4 [1.6] | 11.6 [1.6] |
| (Range) | (12-15.4) | (12.1-15.3) | (6.4-11) | (10.1-14.2) |
| Platelets (x10<sup>3</sup> /μl) |  |  |  |  |
| Average [SD] | 404.3 [85.9] | 300.0 [124.3] | 294.9 [157.2] | 153.5 [28.9] |
| (Range) | (223-468) | (179-461) | (170-608) | (113-183) |
| International Normalized Ratio | 1.3 [0.1] | 1.2 [0.1] | 1.4 [0.0] | NC |
| Admission D-Dimer (ng/mL) | 354 | NC | 699 | NC |

Average and range of laboratory blood tests from COVID-19 patients during their stay on ICU. Abbreviations: Standard Deviation (SD), Not collected (NC)

Enzymatic activity data are reported in **Figure 1** (coagulation enzymes) and in **Figure 2** (trypsin-like enzymes and exopeptidases). **Figure 1** shows that plasmin activity remained stable over time and between patients, serving as internal control of enzymatic activity. Thrombin and Factor IX activity showed an increasing trend over the first five days of ICU admission followed by decrease. Factor X activity decreased over time in Patients 1 and 2 compared to day 2 and increased in Patients 3 and 4. The main finding was the marked increase in Factor VII activity and tryptic-like activity, particularly at a terminal Val-Arg (**Figure 2**) in Patient 3 on day 7, when bacterial superinfection was diagnosed.

**Figure 1.**
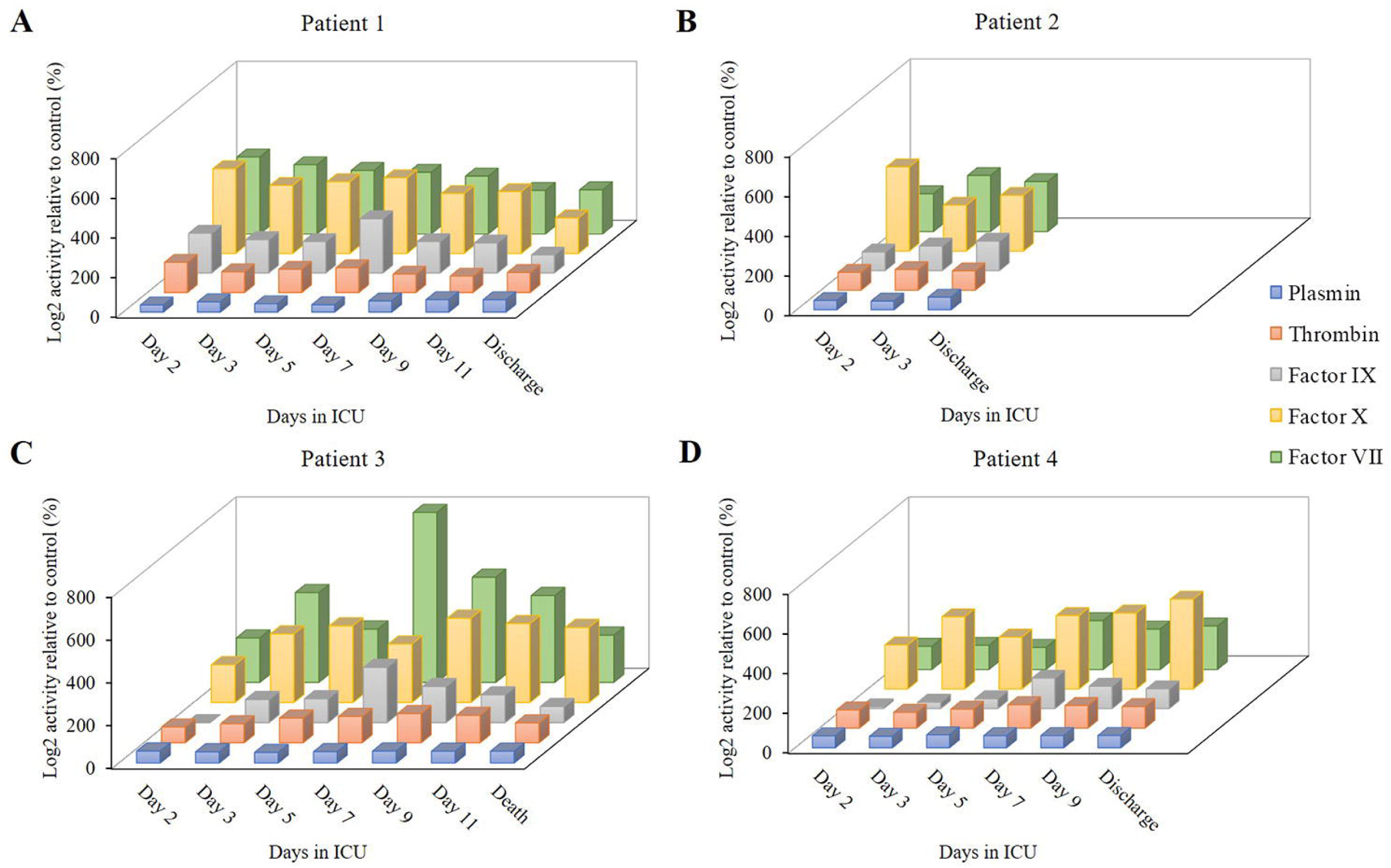
Clotting enzyme activity during the ICU stay of the four patients enrolled in the study.

**Figure 2.**
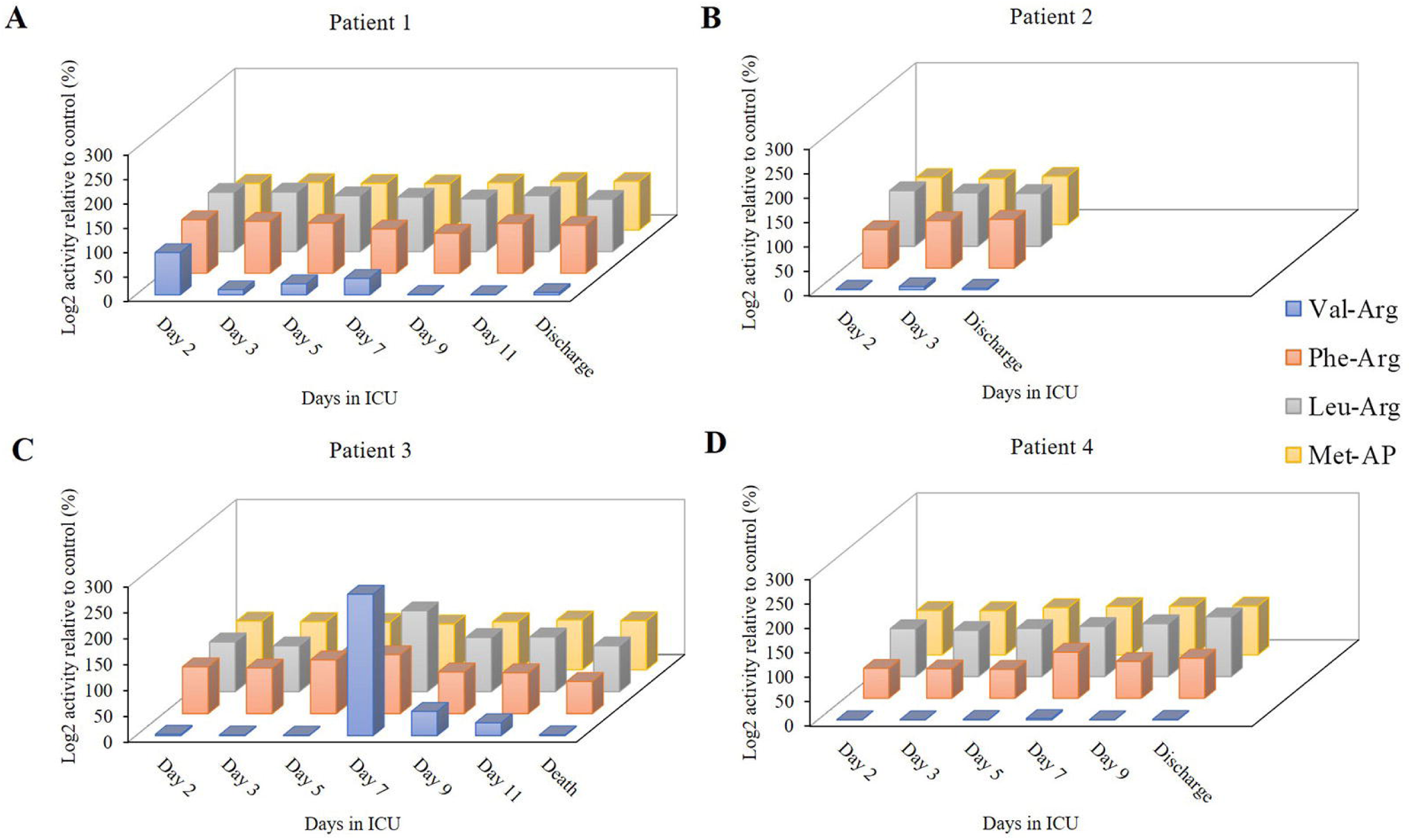
Trypsin-like and amino-peptidase activity during the ICU stay of the four patients enrolled in the study.

**Figure 2** also shows static amino- peptidase activity (Met-AP (Vivitide®, Gardner, MA)), serving as a de facto negative control for the assay.

Patient 3 exhibited a reduction in several plasma coagulation proteins such as coagulation factors V and XI on day 7 and thereafter, and a concomitant increase in the expression of fibrinogen, plasma kallikrein and von Willebrand factor (**Figure 3A**). Overall, there were mild changes over time in plasma from Patient 3 in the abundance of proteins involved in the immune and inflammatory response (**Figure 3B**), as shown by diverging trends of increase and decrease of the complement factors. Interestingly, starting on day 7 ficolin-1 and VEGF 2 were increased in Patient 3, while the trend was less apparent and characterized by some variability in the other patients. C-reactive protein showed a progressively decreasing trend in expression over time in all four patients. A general trend towards a reduction in the expression of enzymatic inhibitors, in particular of endogenous trypsin inhibitors, was observed in Patient 3, (**Figure 3C**). Thrombin inhibitors such as antithrombin-III and heparin cofactor II were also characterized by lower abundance in Patient 3 compared to the survivors at all time points.

**Figure 3.**
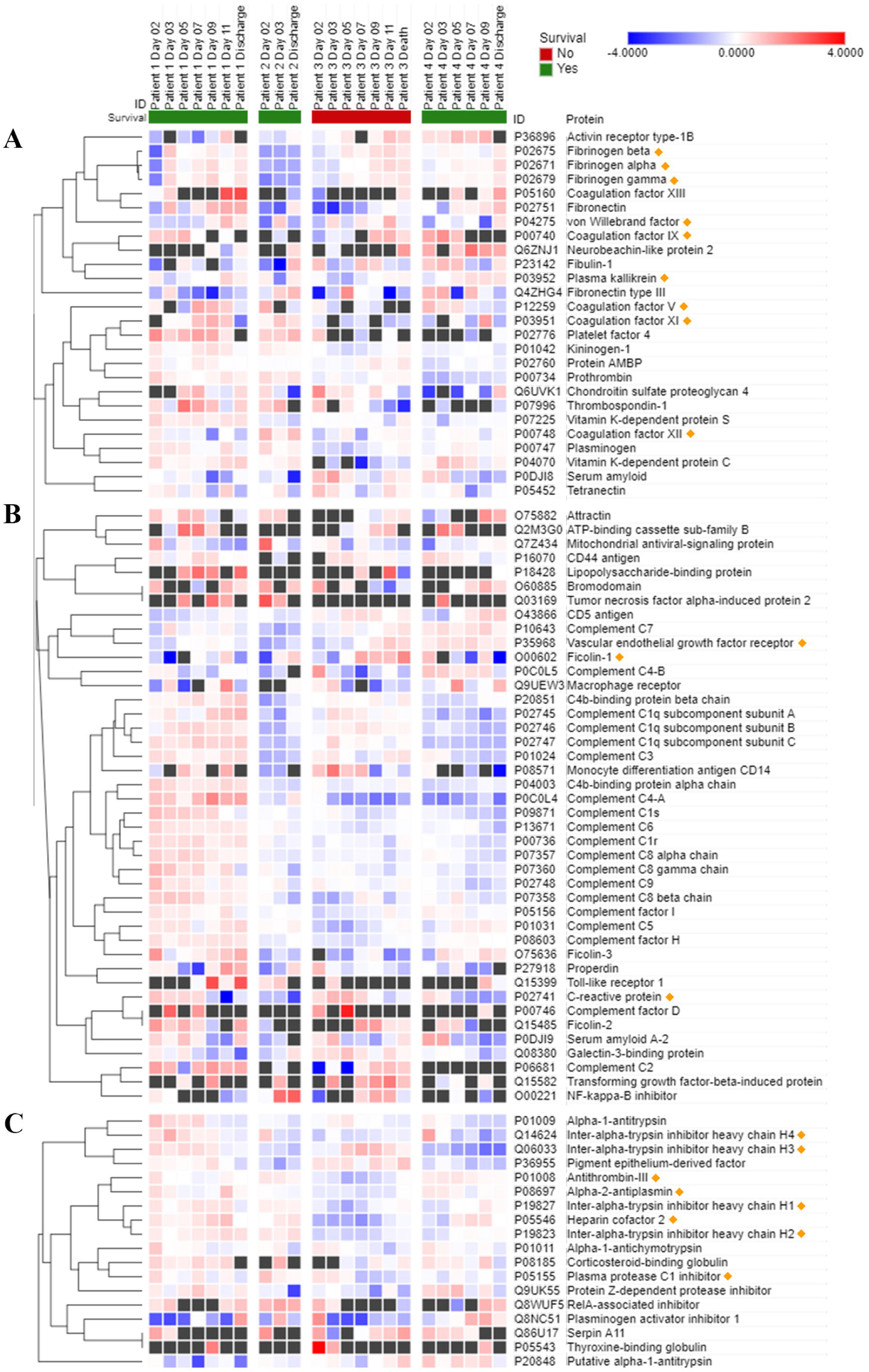
Heatmaps of the most significant proteins involved in coagulation (A), immune or inflammatory response (B), and endogenous enzymatic inhibition (C) detected in COVID-19 patient plasma. Data are presented as Log2 ratio of the averages. Rows of identified proteins were hierarchically clustered with the one minus Pearson’s correlation metric and average linkage method for each protein group. Columns were sorted by patient and ordered by day of sample collection. The heatmaps were obtained using the visualization software Morpheus (https://software.broadinstitute.org/morpheus/).

The proteins characterized by the most evident trends (over time and between patients) are highlighted in the heat maps in Figure 3 with a diamond sign next to the protein name. For the complete proteome profile, please see **Supplementary File 2**.

The sum of the intensities of all the peptide fragments detected at each time point from a total of 233 source proteins suggests marked proteolytic activity in Patient 3 on day 7 (**Figure 4A**). This ongoing protein degradation was particularly evident in fibrinogen-α and fibrinogen-β (**Figure 4B and 4C**). Of note, peptides derived from α2-antiplasmin had a characteristic trend in Patients 1, 3 and 4, with a progressive increase until day 5, followed by an abrupt reduction and a progressive increase thereafter. **Figure 4E** shows the unique patterns of protein cleavage for fibrinogen-α obtained by Peptigram analysis and the size and location of the fragments that suggest that the enhanced proteolytic activity seen on day 7 in Patient 3 increases the amount of circulating fibrinopeptide A (FPA). The link to the complete peptidome profile on Peptigram can be found in **Supplementary File 1**.

**Figure 4.**
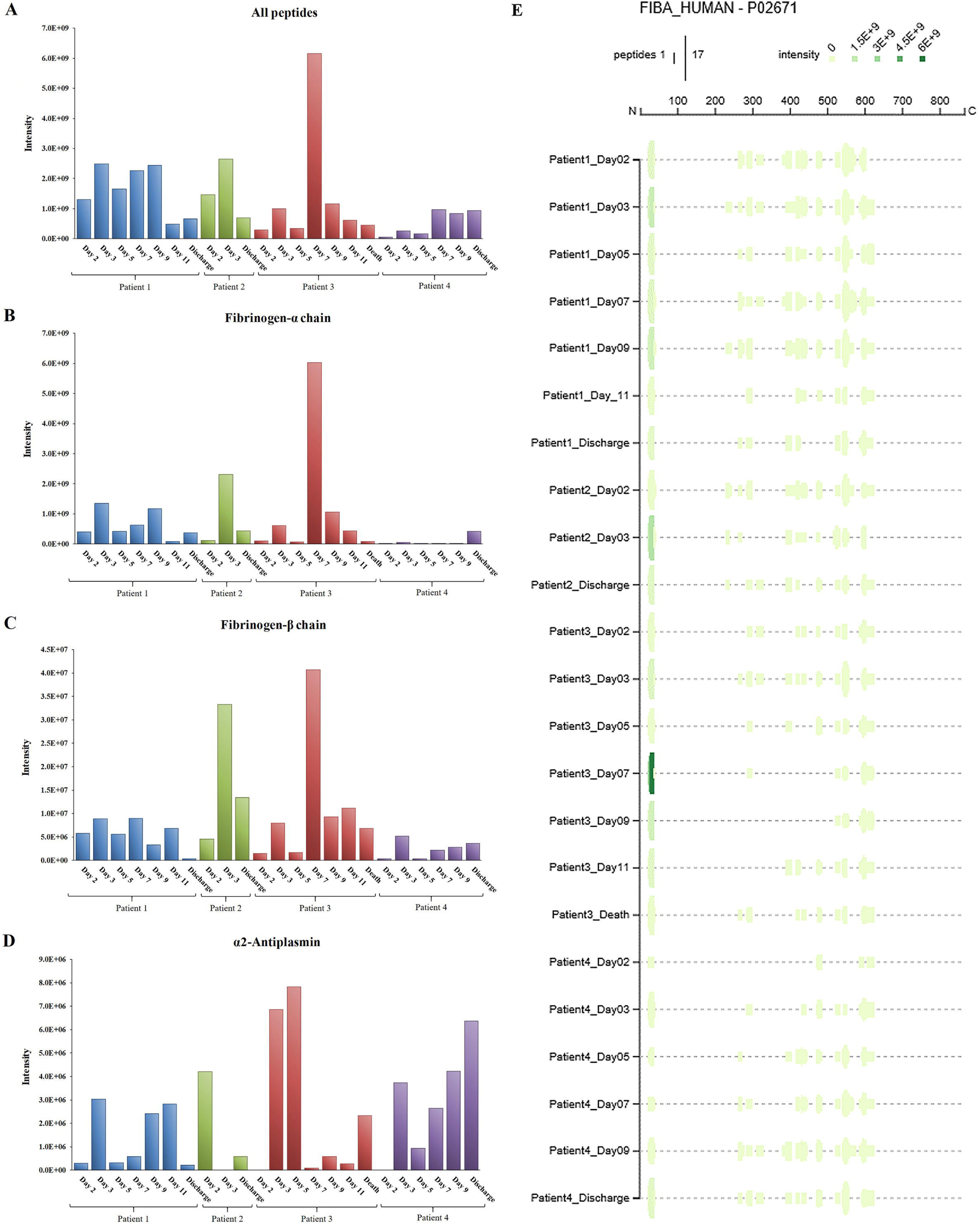
Total intensity as the sum of the intensities of the peptides detected in COVID-19 patient plasma: a) from the whole proteome (A); b) from fibrinogen-α (B); c) from fibrinogen-β (C); d) from alpha-2-antiplasmin (D). Peptigram representation of the cleavage pattern of fibrinogen-α in the four patients at each time point (E).

## 4. Discussion

Omics approaches (metabolomics, proteomics, peptidomics, transcriptomics, genomics) to study disease, including acute illness, have become increasingly popular due to the unique multiscale insights they provide that can result in better targeted treatments (12,14,18-23). Several contributions have already been made to the field of COVID-19 by proteomics (24-27).

We have previously leveraged -omics analyses to test the hypothesis that dysregulated systemic proteolysis occurs in circulatory shock using a peptidomic-based assay, which estimates proteolysis by assessing the abundance and count of circulating peptides (13,14). Analysis of the plasma peptidome by this method showed an association between enhanced proteolysis and death (14) in septic shock caused by bacterial infection. Therefore, we investigated whether a similar association may also be found in viral sepsis caused by COVID-19 infection with bacterial superinfection developing over the course of the ICU stay. To better characterize and quantify the dynamic aspect of systemic intravascular proteolysis, proteolytic activity generated by proteases that are known to be activated in acute illness, such as clotting factors and other serine proteases were measured.

Protease activity patterns showed a marked increase of Factor VII and Factor IX activity on day 7 in the patient who died of superinfection. In particular, the spike in Factor VII activity was unique amongst our sample cohort and represents an 80-fold increase in activity relative to that found in healthy control plasma. Further, the same trend in activity was also found in two substrates identified by the C-terminus cleavage sites Val-Arg and Leu-Arg, representative of tryptic-like proteolytic activity. Although the activity of all four substrates (Factor VII, Factor IX, Val-Arg, Leu-Arg) tended to decrease after the initial increase, the peak on day 7 in Patient 3 coincided with the diagnosis of bacterial superinfection, and with an increase in peptide abundance, particularly from fibrinogen.

Most fibrinogen-derived peptides in Patient 3 on day 7 originated from the Fibrinopeptide A (FPA) domain, located at the N-terminal region of the fibrinogen Aα chain (Aα 20-35), which is normally generated by thrombin cleavage of fibrinogen upon activation of the clotting cascade. FPA is a known biomarker of the activation of the coagulation system and was shown to be related to elevated inflammation in sepsis, including COVID-19-induced sepsis (28). Although thrombin was not shown to be overactivated, other serine proteases may contribute to fibrinogen cleavage at typical thrombin cleaving sites. Further, the underexpression of antithrombin-III and heparin cofactor II also hint at a decreased capacity to regulate thrombin and other protease activity, regardless of thrombin expression and activity levels. Consistent with our prior findings on enhanced proteolysis over time in patients who died of septic shock (14), these findings may indicate a possible relationship between a surge in proteolytic activity in concert with impairment of the coagulation cascade and worsening overall clinical course after development of bacterial infection following the initial progression of COVID-19 infection. This was apparent clinically in Patient 3 who had been on either phenylephrine or norepinephrine (but not both) for hypotension from admission to the ICU, and on day 7 necessitated the addition of two simultaneous pressor agents (norepinephrine and vasopressin) to maintain adequate hemodynamics. This pressor requirement subsequently increased the following day to three simultaneous pressor agents (norepinephrine, phenylephrine, and vasopressin) before later subsiding.

As COVID-19 infection has a strong association with coagulopathy (29,30), we focused our study on the activity of enzymes involved in the clotting cascade and other trypsin-like proteases, which could be characterized by a similar array of target proteins, including but not limited to coagulation enzymes and proteins such as fibrinogen. Thromboembolism and reports of microvascular thrombi akin to a disseminated intravascular coagulation (DIC) picture have been a common reported cause of death in patients admitted to the ICU with severe COVID-19. Although the cause of death in Patient 3 was classified as septic shock, the possible association between enhanced proteolytic activity, bacterial superinfection and disease severity is nonetheless of interest and could point to i) the presence of additional pathological mechanisms (i.e., proteolysis) that contribute to poor outcomes; ii) the relationship between enzymatic activity patterns and bacterial superinfection that causes further illness, in addition to that already seen with precedent viral infection.

The association between disease severity and -omics patterns is also supported by the measurements of other proteins that play a role in immunity and inflammation. For instance, the trends in expression of ficolin-1 and VEGFR2, and to some extent C-reactive protein (which showed a decrease over time in all four patients, much milder in Patient 3 though) correlate with the exacerbation of the inflammatory state of Patient 3 following diagnosis of bacterial superinfection. Ficolin-1 is a known activator of the complement system and innate immunity through recognition of pathogen-associated molecular patterns (PAMPs) (31,32), while VEGFR2 has been related to COVID-19 progression (33) as also suggested by the hypothesis that the VEGF receptor activator (PMC18457) may serve as a biomarker for detecting the progression of COVID-19 (34). In fact, the patient with the least severe course of disease (Patient 2) had the lowest VEGFR2 intensity and, importantly, was the only patient who was not diagnosed with ARDS. Patient 4, on the other hand, had rheumatoid arthritis and subsequently greater VEGFR2 intensity. These observations highlight that the complexity of the interplay between baseline comorbidities, severity of the disease and variability of the features of immune and inflammatory responses prevents from interpreting conclusively the expression trends of complement factors and warrants further research in the future.

There are some limitations of this study that need to be acknowledged. First, regarding the clinically (obtained from the patient record) derived data, the low sample size of the studied patient population and non-uniformity in the timing of some laboratory results hinders the ability to make stronger inferences than are made here; the robustness of the presented results would doubtlessly have been strengthened with a larger sample size and fewer missing data. However, even with this very limited sample size we were able to make some cautious conclusions as to new hypotheses of molecular mechanisms that might otherwise not be recognized with a larger patient population. Second, regarding data drop-out, this was a real-world observational study, and the clinical laboratory measurements and values reflect patient management of severely ill COVID+ patients in the ICU. Third, in our analytical protocol, the mass spectrometry approach was necessarily untargeted; if specific protein systems and products of their degradation were targeted, it is possible that the potential dysfunction in these systems (e.g., coagulation, complement, etc.) could be highlighted in greater detail. Finally, when comparing and integrating proteomics and peptidomics data, the availability of transcriptomics data, which were not part of the experimental design of this observational study, could add information on protein regulation, and help understand to a greater depth the impact of abnormal proteolysis and protein synthesis, breakdown, and turnover.

In conclusion, we demonstrate here that the analysis of the circulating proteome and peptidome, coupled with quantitation of enzymatic activity in plasma of COVID-19 patients, provides additional data about the progression of the disease. Our approach could prove particularly useful for achieving a more comprehensive description of the impairment of the activation and regulation of coagulation, a complex, protease-mediated system which plays a critical role in the management of acute patients in the ICU. The association between proteolytic activity and protease expression patterns achieved by mass spectrometry investigation of plasma and bacterial superinfection in COVID-19 may also contribute to anticipate the signs of possible superinfection and aid in the early diagnosis and delivery of a timely therapy.

## Data Availability

Data are available via ProteomeXchange with identifier PXD029181

