## Supplemental file 1 for "Dysregulation of circulating protease activity in Covid-19-associated superinfection"

### *Supplementary file 1*

#### 1 Enzymatic Activity Protocol

The peptide sequences used in this assay are the following:

z-Arg-Arg-Leu-Arg-AMC (Leu-Arg);

N-Benzoyl-Phe-Val-Arg-AMC (Val-Arg);

z-Phe-Arg-AMC (Phe-Arg);

Pro-Asp-Phe-Tyr-AMC (Phe-Tyr);

Met-AMC (MetAP);

z-Gly-Gly-Arg-AMC (Thrombin);

Boc-Val-Pro-Arg-AMC (Factor VII);

Boc-Glu(OBzl)-Gly-Arg-AMC (Factor IX);

Boc-Ile-Glu-Gly-Arg-AMC (Factor X);

H-D-Ala-Leu-Lys-AMC (Plasmin).

#### 2 Proteomics Protocol

##### 2.1 Protein Purification and Digestion

2µL of plasma from COVID-19 patients was mixed into 100µL of 6M guanidine solution and boiled twice for 5 minutes each. After allowing the solution to cool, 950µL of methanol was added, vortexed, and incubated for 30 minutes at room temperature. The plasma solution was centrifuged at 13,000g for 20 minutes at 4°C. The supernatant was discarded before introducing 8M urea in 200mM ammonium bicarbonate (AmmBic) to the protein precipitate. The sample was vortexed and then mixed at 1300rpm for 30 minutes at 37°C. A solution of 4µL 0.5M tris(2-carboxyethyl) phosphine (TCEP) and 6µL 8M urea was added to the sample and mixed again at 1300rpm for 30 minutes at 37°C, followed by overnight freezing at -20°C. The frozen sample was warmed up to 37°C for 30 minutes with constant mixing. 10µL of chloroacetamide solution was added and mixed for 30 minutes at 37°C. 50µL of the sample was combined with 50µL of 50mM AmmBic/0.5µg LysC and was incubated at 37°C for 4 hours. 1µg of trypsin in 100µL of 50mM AmmBic was added to the sample for digestion and incubated at 37°C overnight. On the next day, the solution was warmed to 42°C for 1 hour. 10µL of 75% formic acid (FA) was added to the sample and vortexed. Solid-phase extraction was then performed on the peptide products using organic solvents and a polymeric reversed-phase column (8B-S100-UBJ, Strata-X). The following steps were executed in order: 100%

acetonitrile (ACN) activation, 0.2% Formic Acid (FA) equilibration, column separation, 0.2% FA wash, and 40% ACN elution, drying, resuspension in 0.5% FA/5% ACN. 0.5µg of total peptides, determined by BCA assay, was used per run.

#### 2.2 Proteomics Mass Spectrometry Parameters

The set mass spectrometer parameters include: MS1 survey scan via Orbitrap detector and quadrupole filtration (400-1500 m/z mass range, 2400V spray voltage, 60,000 resolution, 290°C ion transfer tube, automatic gain control target of 400,000, 50ms maximum injection time), data-dependent scans (+2-5 charged state ion inclusion, 20 second exclusion time, HCD collision energy of 30% on selected ions of minimal intensities of 50,000), and fragment mass analysis (turbo scan rate of ion trap, automatic gain control target of 5,000, first mass 100 m/z, 35ms maximum injection time).

#### 2.3 Proteomics Search Parameters

PEAKS Studio 8.5 (Bioinformatics Solutions, Inc.) search engine was used with the following parameters:

Parent Mass Error Tolerance: 15.0 ppm
Fragment Mass Error Tolerance: 0.4 Da
Precursore Mass Search Type: Monoisotropic
Enzyme: Trypsin
Max Missed Cleavages: 3
Non-Specific Cleave: Both
Fixed Modifications: Carbamidomethylation: 57.02
Variable Modifications: Oxidation (HW) 15.99, Oxidation (M): 15.99, Phosphorylation (STY): 79.97, Deamidation (NQ): 0.98, Acetylation (K): 4.01, Methylation (KR): 14.02
Max Variable PTM Per Peptide: 3
Database: Human Uniprot
Contamination Database: Contaminants
Searched Entry: 7000
FDR Estimation: Enabled
De Novo Score (ALC%) Threshold: 15

### 3 Peptidomics Protocol

#### 3.1 Peptide Extraction and Preparation

100µL of plasma from COVID-19 patients was combined with 900µL methanol and vortexed. The samples sat at room temperature for 30 minutes before centrifugation at 12,000g for 10 minutes. 500µL of the supernatant was collected and dried using a SpeedVac concentrator (Thermo Scientific). The samples were hydrolyzed in a 0.5mL solution of 0.5% FA and 5% ACN. C18 solid-phase extraction (WAT054955 Sep-Pak C18 Vac Cartridge, Waters Corporation) was conducted following the manufacturer's protocol, except for the substitution of 40% ACN as the elution solution. The eluents were dried and stored until mass spectrometry analysis.

#### 3.2 Peptidomics Mass Spectrometry Parameters

The set mass spectrometer parameters include: MS1 survey scan via Orbitrap detector and quadrupole filtration (400-1500 m/z mass range, 2400V spray voltage, 60,000 resolution, 285°C ion transfer tube, automatic gain control target of 400,000, 50ms maximum injection time), data-dependent scans (+2-5 charged state ion inclusion, 5 second exclusion time, HCD collision energy of 30% on selected ions of minimal intensities of 50,000), and fragment mass analysis (turbo scan rate of ion trap, automatic gain control target of 5,000, first mass 100 m/z, 35ms maximum injection time).

##### 3.3 Peptigram

The full peptidome analyzes can be found on the following link:

http://bioware.ucd.ie/peptigram/job/7jbnkglrh290
