## Supplementary figures and images for "Dysregulation of circulating protease activity in Covid-19-associated superinfection"

### Supplemental file 2

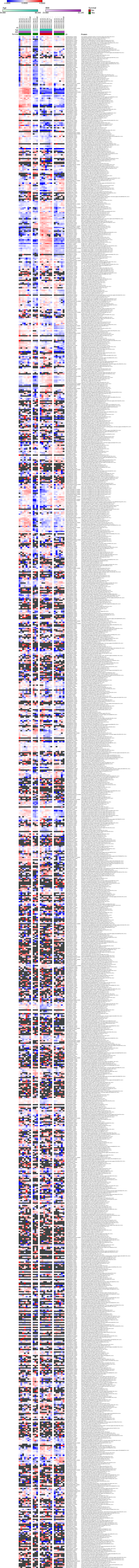
